# COVID-19 patient accounts of illness severity, treatments and lasting symptoms

**DOI:** 10.1101/2021.05.26.21257743

**Authors:** Moriah E. Thomason, Denise Werchan, Cassandra L. Hendrix

## Abstract

First-person accounts of COVID-19 illness and treatment complement and enrich data derived from electronic medical or public health records. With patient-reported data, it is uniquely possible to ascertain in-depth contextual information as well as behavioral and emotional responses to illness. The Novel Coronavirus Illness Patient Report (NCIPR) dataset includes complete survey responses from 1,592 confirmed COVID-19 patients ages 18 to 98. NCIPR survey questions address symptoms, medical complications, home and hospital treatments, lasting effects, anxiety about illness, employment impacts, quarantine behaviors, vaccine-related behaviors and effects, and illness of other family/household members. Additional questions address financial security, perceived discrimination, pandemic impacts (relationship, social, stress, sleep), health history, and coping strategies. Detailed patient reports of illness, environment, and psychosocial impact, proximal to timing of infection and considerate of demographic variation, is meaningful for understanding pandemic-related public health from the perspective of those that contracted the disease.

## Background & Summary

Major discoveries about COVID-19 illness, susceptibility, transmission, and human behavior have been unearthed through utilization of rich medical and public digital record systems. Chasms in health inequity have been revealed.[1-3] Discrete spatiotemporal patterns of public health behavior have been characterized.[4, 5] Individual- and community-level risk factors underlying local transmission of COVID-19 have been identified.[6, 7] Examination of internet searches has revealed that (i) information flow about COVID-19 is inversely relates to positive cases [8] and (ii) there have been population-level shifts over the course of the pandemic from searches pertaining to activity/fitness to more sedentary activities and dietary supplements.[9] Overall, publicly available large-scale databases are tremendously powerful sources of information that have been rapidly deployed to identify crucial determinants of health, aspects of transmission, and core human behavioral adaptations in the context of COVID-19.

The challenge, however, is that these studies are limited by the constraints of electronic fields that serve narrow functions and lack nuance that is intrinsic of individual human stories. As a specific example, electronic medical records (EMR) systems include lists of symptoms, physical examination and laboratory results, treatments, diagnoses and limited demographic information. EMR are not intended to capture information about patient perceptions, and yet we know that the subjective experiences and outcomes of one person to the next are variable and also predictive of future health.[10, 11]. Circumstances of illness occur in diverse socioemotional contexts and relate to other events occurring in an individual’s life. The goal of NCIPR was to aggregate a sizable dataset of first-person accounts of COVID-19 illness, risk, and recovery. NCIPR survey data can be used to weave individual strands of history, environment, perspective, and health together to make new discoveries that will compliment and enrich knowledge about COVID-19 that has been derived from medical and public digital record systems. Further, the NCIPR dataset contains measures of lasting secondary effects of COVID-19 infection that are not readily available in medical record systems.

The NCIPR survey was developed in November 2020 and published in the U.S. National Library of Medicine (NLM) Disaster Management Resources (id:24224) as well as the Open Science Framework (https://osf.io/82rkj/) in early March 2021. Patients with COVID-19 diagnoses were identified from within the New York University Langone Medical Center (NYU Langone) EMR system. Data extraction occurred on February 23, 2021. The full workflow is depicted in **Figure 1**, including record extraction, invitation, online consent, and resulting dataset. Two waves of recruitment invitations were implemented, occurring on February 23 and March 29, 2021. Between waves, four new questions were added to gather additional data on lasting symptom complaints, including duration of symptoms, categories of mood symptoms, and two questions about lasting cognitive complaints. Additionally, five questions were added about blood type, height and weight, history of tonsillectomy and the Macarthur Ladder.[12] The survey was closed to potential respondents on April 7, 2021.

**Fig 1.**
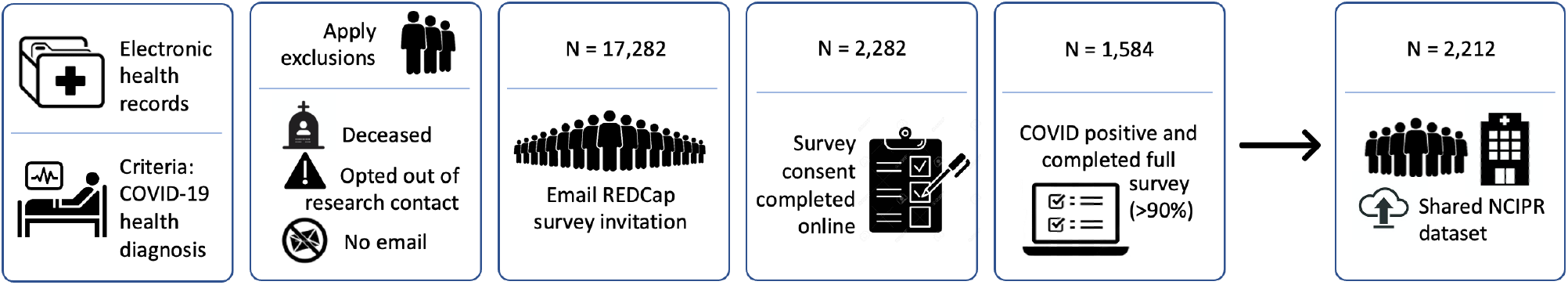
A schematic overview of the study design and data collection workflow.

The primary goal motivating collection of the New York NCIPR dataset was to obtain a comprehensive record of the subjective experiences of those ill with COVID-19, proximal to the time of illness. Along with this, we addressed a number of specific questions percolating across scientific, media and word-of-mouth channels. For example, there have been anecdotal accounts about individuals with pets becoming less ill, about unexpected side effects (e.g., hair loss), about lasting illness sequalae, about underlying vulnerabilities making certain individuals more susceptible. The NCIPR dataset can be used to address a large number of questions that remain unanswered about COVID illness, about human behavior, and about environmental determinants of health. Rapid placement of the data in the public domain better assures that investigation of these and other topics will commence quickly and will be rapidly communicated to wide audiences.

## Methods

### Survey Design

The NCIPR survey was developed to assess COVID-19 symptoms, medical complications, home and hospital treatments, lasting effects, anxiety about illness, employment impacts, quarantine behaviors, vaccine-related behaviors and effects, and illness of other family/household members. The NCIPR also includes questions that address age, financial security, perceived discrimination, pandemic impacts (relationship, social, stress, sleep), health history, and behavioral coping strategies. A subset of questions were adapted from established Common Data Elements for mental health (NLM Disaster Management Resources COVID-19 and Perinatal Experiences (COPE) questionnaire, id:24224; https://osf.io/82rkj/, from the Williams Perceived Discrimination Scale,[13] and from the Fletcher measure of Perceived Relationship Quality.[14] **Table 1** provides a summary of domains covered by the full NCIPR survey.

**Table 1.** Summary of measurement domains assessed by the NCIPR Survey. Data across these domains is contained within the New York NCIPR dataset

| Domain | Topics addressed |
| --- | --- |
| COVID-19 illness | Date of illness; symptoms experienced; exposure; length of illness; fever; perceived severity |
| COVID-19 treatments | Hospitalization; ICU admission; medical treatments; at-home treatments; medications; imaging |
| COVID-19 testing | Timing; type; facility; symptoms present |
| COVID-19 impacts | Loss of income; time off work; quarantine behaviors; perceived life disruption; change in social support, sleep, energy levels, stress, relationship satisfaction |
| COVID-19 perceptions | Anxiety about illness; satisfaction with medical care; opinion about when things will go back to normal |
| COVID-19 lasting effects | Length of symptoms; types of symptoms; specific mood disturbances; specific cognitive disturbances; estimate of time to full return to health |
| Physical characteristics | Age; height; weight; blood type |
| Health characteristics | Preexisting health conditions; prior substance use and mental health treatment; current stress level; prior tonsillectomy |
| Vaccine information and attitudes | Date received; manufacturer; side effects; if not vaccinated, attitudes about vaccination for self and/or children; plans to relax COVID-19 safety behaviors after vaccination; if breastfeeding, attitudes about vaccination and infant side effects |
| Demographic and financial context | Gender identity; employment (self and partner); income change due to COVID-19; stability of housing; public assistance; medical insurance; satisfaction with financial situation; MacArthur Scale of subjective social status |
| Home environment | Pets; number of individuals living in home; number of household members that became ill; number of bedrooms in home |
| Patient behavior | COVID safety behavior; coping strategies; drug, nicotine and alcohol use; exercise; use of meditation/mindfulness; religious practices; family/friend support; screen use; social media use |
| Perceived discrimination | Amount; kind; distress |

*The NCIPR questionnaire also includes questions about child ages, breastfeeding, education, race/ethnicity, income, number of bedrooms in home, utilization of public assistance, and preferred medical health system. For data release and compliance with regulation on indirect identifiers and patient confidentiality, these are removed from released data, as described below.*

### Ethical approval

The research protocol for this study was approved by the NYU Langone Institutional Review Board (IRB). Only patients that had previously consented to be contacted about research opportunities were eligible for invitation into the study.

### Recruitment and Survey Administration

A search of the NYU Langone Health record system identified all individuals ages 18 and older that had been diagnosed with COVID-19. Individuals (1) with email contact, (2) not deceased, and (3) not designated as having previously opted out of research contact were eligible to participate. After application of these exclusions, 17,282 individuals were sent an email inviting them to participate in a 10 to 15-minute survey. Compensation was entry into an end of week drawing for a $25 Amazon gift card. Study data were collected and managed using REDCap electronic data capture tools hosted at NYU Langone University.[15, 16] The measure was administered in English.

### Sample description

The NCIPR dataset contains data from 2,212 individual respondents. 2,147 of these respondents confirm having been ill with COVID-19 in addition to having COVID-19 diagnosis in their medical record. However, description of illness severity and demographics provided here are restricted to 1,584 cases that passed the Technical Validation steps described in the section below. Timing of COVID-19 illness in the sample reflects peak prevalence rates in March 2020 and January 2021 (**Fig. 2**). Illness severity varied across the sample, as seen in length of illness, fever duration, peak fever, hospitalizations, and in self-reported illness severity ratings (**Fig. 2**). Sample demographic data are provided in **Fig. 3**. Respondent ages range from 18 to 98 years old. Due to a survey administration error described below, complete data are available at a ratio of ∼2:1, females to males.

**Fig 2.**
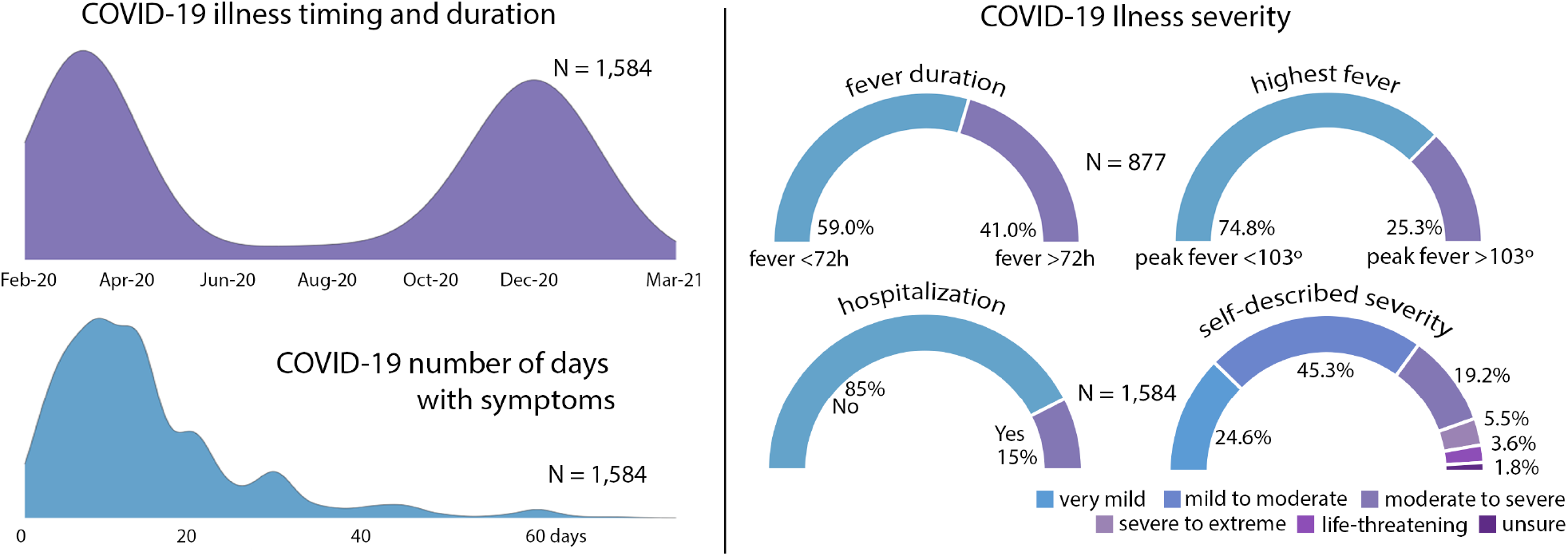
Overview of illness severity in the N = 1,584 COVID-19 quality validated sample

**Fig 3.**
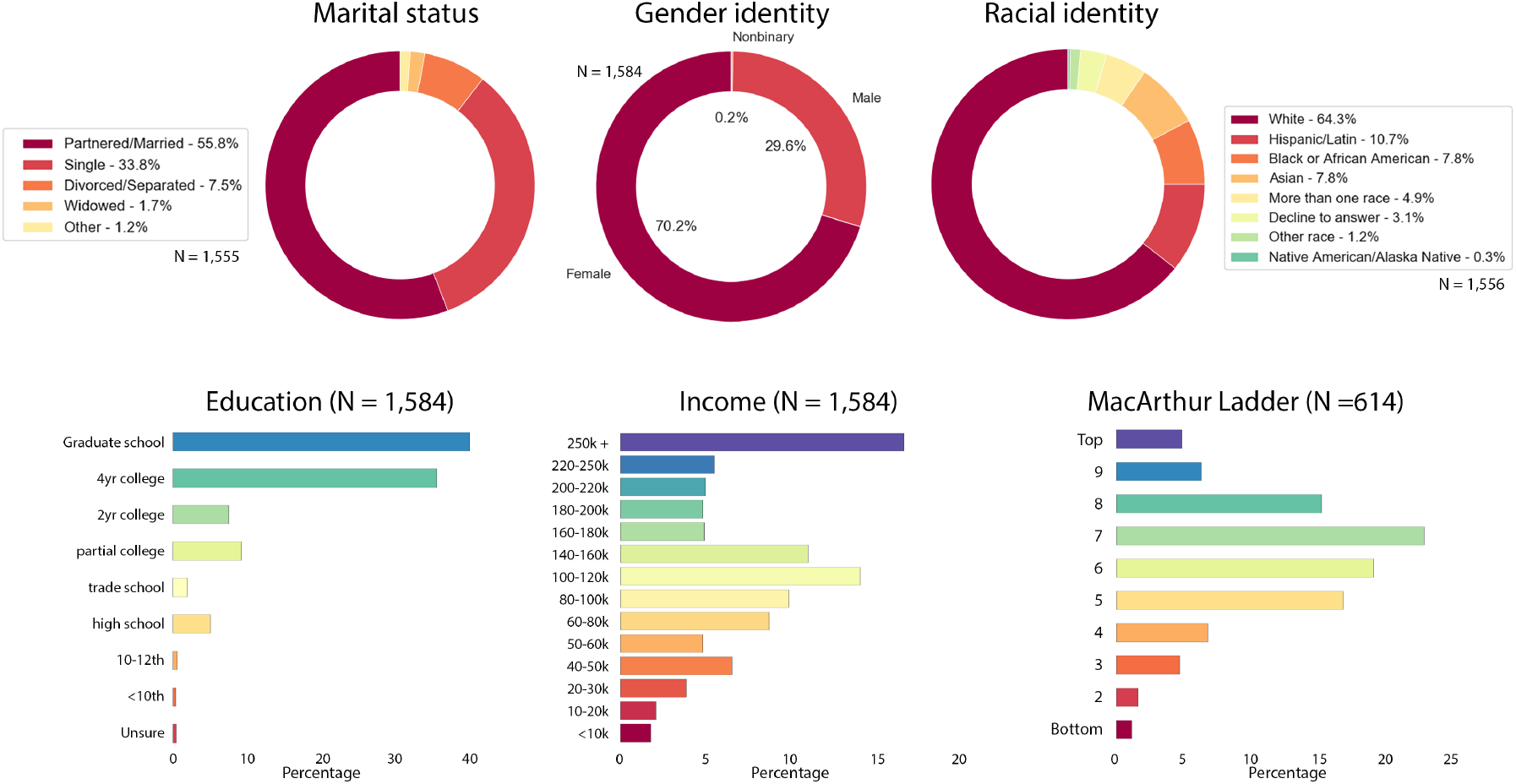
COVID-19 infected sample demographics N = 1,584

### Geo-positioning of COVID-19 survey respondents

Geographical information about survey respondents was derived from a subset of patients (N = 697) that provided consent to future contact within the online consent form. Those that made this selection were asked to provide contact information and zip code data. Zip codes were converted to corresponding Federal Information Processing System (FIPS) codes. The distribution of patient FIPS is displayed in **Fig. 4**. The majority reside in Manhattan, Brooklyn and Long Island. A small number provided zip codes in states other than New York, New Jersey and Connecticut, N = 9.

**Fig 4.**
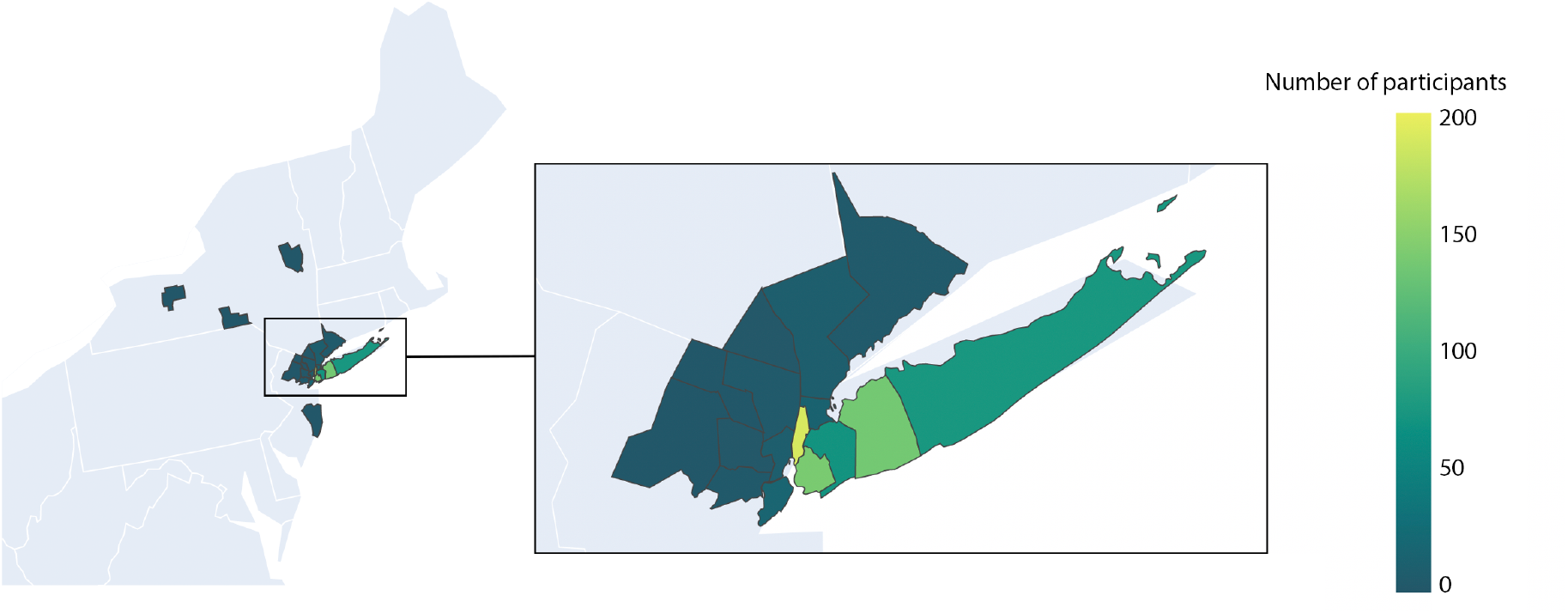
Geographical location of COVID-19 patient survey respondents

### Data Records

The dataset resulting from the NCIPR survey is stored in a CSV format via the https://osf.io/82rkj/ open access platform. Each row represents one respondent and each column represents a variable. The file includes every survey respondent except for those who completed the consent form only (N=68). Date of birth was converted to age in years, variable name [age_calculated]. A second variable, [db_52], is the age in years provided by the participant. Inclusion of both age-related data fields was intentional, as this provides a means of data validation, described in more detail below. Ordering of the variables in the CSV files reflects the order in which items were administered. During data preparation and validation, 10 variables were added to aid in future data processing. **Table 2** summarizes the variables that were added to the raw data set during quality assessment and data validation.

**Table 2.** Summary of variables added to dataset during preparation and validation steps

| Variable name | Variable definition |
| --- | --- |
| which_NCIPR | (1) NCIPR wave 1 February 23, 2021; (2) NCIPR wave 2 March 31, 2021 |
| complete_binary | (0) incomplete (n=483); (1) complete (n=1,729) |
| why_incomplete | (1) complete (n=1,729); (2) survey administration error (n=338); (3) incomplete survey (n=145) |
| covid_self_report | (0) report no prior COVID-19 illness (n=65); (1) confirm COVID-19 prior illness (n=2,147) |
| DOB_age_out_of_range | (0) date of birth age = 18-100 years (n=2,157); (1) date of birth age = <18 or age >100 (n=55) |
| COVID_date_out_of_range | (0) Feb 2020 - March 2021 (n=2,192); (1) dates in range not selected (n=20) |
| quality_check_flag | (0) none (n=1,857); (1) $\geq 1$ implausible response (e.g., 6'20" tall) (n=4); (2) $\geq 1$ inconsistent response (What is your current age? [db_52] ≠ reported date of birth +/- one year) (n=68); (3) inconclusive (e.g., age or DOB response not provided) (n=283) |
| data_correction | (0) no correction; (1) typo in age or height; original data unchanged but [quality_check_flag] changed to '0' (n=7) |
| excluded_sample | (0) included (n=1,584); exclusions filtered in the following order: (1) incomplete (n=145); (2) survey admin error (n=338); (3) [covid_self_report] = '0' (n=65); (4) DOB provided out of range (n=46); (5) [quality_check_flag] = '1' or '2' (n=19); (6) COVID-19 illness date inconclusive (n=15) |
| age_calculated | Participant reported date of birth [db_2] converted to age in years |

Additional files released with the primary dataset (.csv) are: (a) the NCIPR questionnaire (.pdf), (b) the NCIPR demographics form (.pdf), (c) the REDCap instrument files (.zip), and (d) the variable definition file (.csv). All are accessible via the Open Science Framework (OSF) open access platform. The questionnaires include response options for each question along with the coding used for each variable. The REDCap files shared via https://osf.io/82rkj/ include the nine additional questions added between recruitment waves. As is evident in ordering of variables in the CSV file, the NCIPR covid illness survey was administered prior to the NCIPR demographics survey.

### Survey administration error

An error in branching logic was identified after the first wave of data collection, such that respondents who did not endorse being female were not offered the majority of questions about COVID illness. This error resulted in a systemic loss of data in 322 male and self-describing gendered participants for COVID illness questions. These cases are included in the shared dataset and are designated as such, as referenced in **Table 2** and in the added variable [why_incomplete] = 2.

### Removal of indirect identifiers

Confidentiality and anonymity are key ethical considerations when publishing or sharing data relating to individuals.[17] Indirect identifiers removed from the dataset are indicated in the data variable definition file available on OSF, https://osf.io/82rkj/. Indirect identifiers removed include race, ethnicity, income, education, DOB, ages of children, number of bedrooms in home, breastfeeding questions, use of public assistance, number of adults and children in home, and affiliation with NYU hospital system. Further, all dates in the dataset were converted to Month-Year format (e.g. Mar-21) and individuals age 90 or older were edited to 89+ to disallow potential re-identification.

### Technical Validation

Data assurance and quality checking were performed using R version 4.0.2 and Excel. **Table 2** provides a summary of variables added to the dataset during quality validation steps, inclusive of QA/QC codes assigned to survey respondents. Criterion assessed for determinations about quality of patient responses included isolating implausible and/or inconsistent responses. Patients were flagged [quality_check_flag] as (1) “implausible” if they provided a height feet value greater than 7, or a height inches value greater than 12; (2) “inconsistent” if the self-reported date of birth (DOB) and current age were incongruent (defined as different by >1 year); or (3) “inconclusive” if DOB or age in years was not provided. It was noted that 5 individual respondents gave their full height in inches (e.g., 5.2 was entered as feet and 62 was entered as inches), and 2 participants typed a decimal point before self-reported age in years that matched the date of birth provided (e.g., born in 1997 and provided age .24). For those 7 cases, the [quality_check_flag = 1] was changed to [quality_check_flag = 0] and they were included in the final sample, [final_sample = 1], but the raw data causing the flag was not changed. Patient age was computed based on DOB and inserted as a new variable in the dataset [age_calculated]. Findings from these preparation and validation steps guided selection of a final sample that is coded as [excluded_sample] = ‘0’ in the released data; these are the 1,584 described above as passing technical validation for which group level demographics are provided.

## Data Availability

The dataset resulting from the NCIPR survey is stored in a CSV format via the https://osf.io/82rkj/ open access platform. Each row represents one respondent and each column represents a variable. The file includes every survey respondent except for those who completed the consent form only (N=68). Ordering of the variables in the CSV files reflects the order in which items were administered.

## Acknowledgements

This project was supported by a COVID-19 supplement award to MET from the National Institutes of Health connected to awards R34DA050287 and R01MH126468. The authors thank Autumn Austin, Carly Lenniger, and Amin Majbri for contributions to recruitment and data collection. The authors thank participants who generously shared their time and who expressed interest in helping researchers and clinicians better understand the varied experiences and circumstances surrounding COVID-19 illness and recovery.

## Author contributions

MT designed the experiments. DW and CH analyzed the data. MT contributed materials. MT, DW, and CH wrote the paper.

## Competing interests

The authors declare no competing interests.

## Notes

### Competing Interest Statement

The authors have declared no competing interest.

### Author Declarations

NYU Langone Institutional Review Board (IRB)

